# The role of exercise in limiting progression from liver inflammation and fibrosis to cirrhosis and carcinoma: a systematic review with meta-analysis of human and animal studies

**DOI:** 10.1101/2023.08.17.23294088

**Authors:** E.N. Stanhope, A.E. Drummond, C.T.V. Swain, N. Teoh, G. Farrell, J.K. Vallance, I.M. Lahart, B.M. Lynch

## Abstract

**Background:** Exercise may prevent the progression of liver disease and protect against liver cancer. This review with meta-analysis synthesised the evidence from both human and animal studies to better understand whether exercise has the capacity to (i) promote regression of early fibrosis; (ii) decrease and/or delay progression to cirrhosis; and (iii) progression to carcinoma.

**Methods:** A systematic search was performed to identify studies comprising of humans and animals with liver disease that compared exercise to an inactive or less active control. Outcomes included liver disease regression and progression, and markers of liver function and damage.

**Results:** We found 18 human and 29 animal studies. A single study provided direct evidence that exercise can reverse NAFLD and decrease progression to cirrhosis. Meta-analysis of human studies identified decreases in liver enzymes; ALT (SMD = -0.28, 95%CI = -0.53, -0.03), AST (SMD = -0.12, 95%CI = -0.32, 0.07), GGT (SMD = -0.23, 95%CI = -0.36, -0.10), as well as a small increase in ALP (SMD = 0.23, 95%CI = -0.13, 0.59), and liver triglycerides (SMD = -0.24, 95%CI = -0.66, 0.18). Meta-analysis of animal studies identified decreases in liver enzymes; ALT (SMD = -2.85, 95%CI = -4.55, -1.14), AST (SMD = -2.85, 95%CI = -4.55, -1.14), and liver triglycerides (SMD = -1.36, 95%CI = -2.08, -0.65), liver weight (SMD = -1.94, 95%CI = -2.78, - 1.10), and the NAFLD activity score (SMD = -1.36, 95%CI = -2.08, -0.65).

**Conclusion:** Only one study provided direct evidence that exercise has the capacity to regress early fibrosis, as well as delay the progression to cirrhosis. Several studies, however, indicate that exercise intervention reduce markers of liver function and damage.

## Introduction

Liver disease is a growing concern worldwide with rising incidence rates irrespective of age, sex, region, or ethnicity (1). The global prevalence of non-alcoholic fatty liver disease (NAFLD) has been estimated to be between 20% and 30% in adults (2), of which ∼20% have non-alcoholic steatohepatitis (NASH) (3). The term NAFLD encompasses a continuum of liver pathologies, ranging from non-alcoholic fatty liver (NAFL) to cirrhosis and hepatocellular carcinoma (HCC) (4). Simple steatosis with minimal inflammation (NAFL) is relatively benign and has a much lower risk of disease progression than patients with active inflammation (NASH) (5,6). However, once liver fibrosis is present the risk of disease progression and liver-related complications increases dramatically (7) and the chance of developing HCC is seven times greater than that of patients without liver disease (8).

According to the World Health Organisation (WHO, 2020), the progression from acute hepatitis B virus (HBV) to chronic HBV is relatively rare, affecting less than 5% of cases. People with chronic HBV infection, however, face a notable lifetime risk of developing cirrhosis or liver cancer (15% to 40%) (9). Conversely, the transition from acute hepatitis C virus (HCV) to chronic HCV is more common. This transition is observed in 55% to 85% of people with HCV, with approximately 15% to 30% of these people progressing to cirrhosis. In both the case of HBV and HCV, the prognosis for people who develop liver cirrhosis or HCC is considerably worse (10–12). Given that HCC is the third leading cause of cancer-related deaths worldwide (12), and approximately 90-95% of people with HCC have underlying liver cirrhosis (13), the progression from liver cirrhosis to HCC is a matter of great concern.

The liver is one of only a few organs that can regenerate. Its regeneration capacity, however, is finite. If there is insufficient availability of hepatocytes, liver disease can become irreversible with severe consequences, such as increased sensitivity to noxious substances (encephalopathy), increased risk of gastrointestinal (GI) bleeding (varices and coagulopathy), and decreased bile flow (14).

Lifestyle modification, such as increasing physical activity, may help improve liver disease outcomes. Emerging evidence suggests that increasing physical activity may enhance liver repair, reduce the risk of liver diseases, such as NAFLD and liver cancer, and slow the progression from HBV, HCV, NAFLD, and alcohol-related liver disease to liver cirrhosis and HCC (11,12). Whereas the epidemiological evidence suggests a protective effect of increased physical activity against liver cancer, the experimental evidence remains inconclusive—largely due to factors such as heterogeneity in the assessment of ‘exposure’—which limits support for exercise intervention in treating liver disease (15).

Examining the evidence for the biological plausibility of exercise reducing the risk of liver disease can provide support for causal inference. Preclinical studies indicate that, if hepatic steatosis can be reversed through exercise intervention, then liver inflammation and fibrosis can be resolved (4). Due to the invasive nature of liver biopsy, much of the mechanistic evidence for the effects of exercise on liver disease comes from animal studies. However, in vivo animal studies suggest liver fibrosis is more susceptible to regeneration in animals compared to humans (16), and therefore species-specific liver regeneration differences cannot be ruled out (17). Hence, to ensure translation, human mechanistic studies must also be examined.

Developing a better understanding of how exercise affects intermediate biological processes may provide insight into the best ways to alter exposures for disease prevention. The overarching objective of this review is to improve the understanding of whether exercise has the capacity to (i) promote regression of early fibrosis, (ii) decrease or delay progression to cirrhosis; and (iii) progression to carcinoma.

## Methodology

### Study Design

This was a systematic review and meta-analysis of human and animal studies.

### Search Strategy

Relevant publications were identified using tailored search strategies of MEDLINE (via Ovid) from 1946 - June 27, 2021, EMBASE (via Ovid) from 1974 - June 27, 2021, and SportDiscus (via EBSCO) from 1985 - July 4, 2021. The tailored search strategies used a combination of medical subject headings and free text terms. A version of the search strategy for MEDLINE is presented in Appendix 1. Searches were performed on June 29, 2020, and updated on July 22, 2021.

### Eligibility criteria

Both human and animal studies published in peer-reviewed journals were eligible for inclusion. Only studies published in English were included. Conference abstracts were excluded.

We included human studies with parallel group randomized controlled trials (RCTs) and randomized crossover trials (RXTs) designs, whereas non-randomized interventions and observational studies were excluded due to their lower level of evidence for establishing causal effects. Participants included adults (>18 years old) with liver inflammation (including NAFL and NASH) or fibrosis. Populations with other clinically relevant health conditions (e.g., metabolic syndrome, type II diabetes) were also considered. Acceptable interventions included any exercise intervention that disturbed the bodies homeostasis through muscular activity, whether exclusively concentric, eccentric, or isometric, or a combination thereof (18). We considered studies that offered general dietary recommendations in addition to exercise but excluded studies that combined exercise and dietary interventions (e.g., caloric restriction). We excluded studies if the effects of exercise could not be isolated and if the period of exercise was less than seven days. We included studies that contained at least one control arm that did not receive exercise.

Regarding animal studies, we included parallel randomized controlled trials involving genetic and induced models of liver disease; induction methods included, but were not limited to, exposure to diet and alcohol. We excluded single-group pre-post intervention studies and those lacking a non-exercise period for NAFLD induction or a non-exercise induction period which lasted less than eight weeks, as it is improbable to induce NAFLD within such a short timeframe (19,20). Eligible interventions included any form of exercise as defined by Winter & Fowler (2009). We excluded studies if the effects of exercise could not be isolated (e.g., exercise and caloric restriction). To be considered for inclusion, the period of exercise intervention must have been greater than seven days and studies were required to have at least one control group that did not receive exercise.

We included studies that used invasive or non-invasive procedures to determine whether i) inflammation or fibrosis had reverted to an earlier stage of the disease or resolved; (ii) progressed to cirrhosis; (iii) and/or progressed to carcinoma.

The gold standard for evaluating liver fibrosis, cirrhosis, and carcinoma (cancer) is a liver biopsy, whereby a small tissue sample from the liver is extracted for microscopic examination, enabling the assessment of fibrosis (scarring), cirrhosis (advanced scarring), and the presence of carcinoma or cancerous cells. However, liver biopsy is an invasive procedure that may have some risks associated with it, such as bleeding or infection. As a result, non-invasive methods, including FibroScan (Transient Elastography), blood tests, magnetic resonance elastography (MSE), shear wave elastography (SWE), fibrosis blood panels and other imaging modalities (i.e., CT and MRI) have been developed and were included in our review.

### Selection of studies

Study eligibility was judged against the pre-defined inclusion and exclusion criteria. Two authors (ES, IML) independently screened the titles and abstracts of articles for eligibility. In cases of disagreement the two authors discussed each case and tried to reach a consensus. If a consensus could not be reached a third reviewer was consulted (BL). If inadequate information was available in the title and abstract to determine eligibility it was included for next stage (full text) review. The same two authors (ES, IML) independently reviewed the full text of articles that were included based on the title and abstract screening. In cases where there was disagreement about the eligibility of an article, the two authors aimed to reach a consensus. If a consensus could not be reached a third reviewer (BL) was consulted. The authors were not blind to study authors, journals, or results. A screening log was maintained and the number of excluded studies and their reason for exclusion recorded.

### Data extraction

Data extraction for human studies was performed by ES and IML using a pre-piloted system, whereby one author extracted data into a table, and another validated the data extraction. Data extraction for animal studies was performed in the same way by BL and AD. Extracted data included study details (e.g., authors, year), study design, population (e.g., sample size, demographic information, health status), intervention details (e.g., duration, exercise frequency, intensity, time, type), comparator conditions, outcome details (e.g., definition, assessment method), and statistical measures (e.g., post-intervention means and standard deviation). The Webplot digitiser was used to extract data from studies that exclusively presented their results in graphical form.

### Data synthesis and meta-analysis

Extracted data were summarised and presented descriptively. For outcomes that were consistently defined and reported in at least three studies, random effects meta-analysis was used to estimate the effects of exercise on liver outcomes. Meta-analysis compared post-intervention means and standard deviation between intervention and control groups. When studies had more than one intervention group that met the criteria for inclusion, post-intervention means and standard deviations were combined as per the Cochrane Collaboration manual (21). Meta-analysis effect estimates are presented as standardised mean difference (SMD) and 95% confidence interval (95%CI). Heterogeneity was assessed using the I^2^ statistic. Publication bias was assessed via visual inspection of funnel plots. For animal studies, meta-regression was used to assess the influence of study year, study location (i.e., region), animal type, disease, and disease inducement method on effect estimates. For human studies, meta-regression was used to assess the influence of study year, study region, and disease type on effect estimates. Results of studies not included in the meta-analyses were presented descriptively. All statistical analyses were performed using Stata version 16 (Stata Corporation, College Station, Texas).

## Results

### Identification of studies

Search results are presented in Figure 1. Of the 1,535 records identified via database search, 1,352 titles and abstracts were screened, and 174 full texts were assessed for eligibility, with 60 full texts meeting all criteria for inclusion. Of these, there were 29 interventions in animals and 18 interventions conducted in humans.

### Study characteristics

Study characteristics of human studies are presented in Table 1. Studies were conducted in Asia (animal = 6, human = 4), Australia (human = 1), Europe (animal = 8, human = 6), the Middle East (animal = 3, human = 3), North America (animal = 9, human = 1), or South America (animal = 4, human = 1). A total of 1,976 participants with a median (IQR) sample of 47 (27–135) There was a slightly higher proportion of male compared to female participants across studies (median % = 52% vs. 48%); two studies included only female participants (22,23). The median (IQR) average age was 53 (49–55) years and median (IQR) average BMI was 32 (30–33) kg/m^2^.

**Table 1.** Characteristics of included human studies.

| Study | Location | Randomised design | Diagnosis | Sample N | Female (%) | Follow-up duration (weeks) | Intervention: comparison N | Intervention arm(s) description | Comparison arm(s) description |
| --- | --- | --- | --- | --- | --- | --- | --- | --- | --- |
| Abdelbasset 2020 | Egypt | Controlled | NAFLD | 48 | 43 | 8 | 32:16 | a) MICT<br>b) HIIT | Control |
| Balducci 2015 | Italy | Controlled | T2DM | 606 | 42 | 52 | 303:303 | a) Low-MICT + RT<br>b) Mod-HICT + RT | Standard care |
| Cheng 2017 | China | Controlled | NAFLD | 115 | 77 | 34 | 58:57 | a) MICT<br>b) MICT + diet | a) No intervention<br>b) Diet |
| Hallsworth 2011 | UK | Controlled | NAFLD | 19 | NR | 8 | 11:8 | RT | Control |
| Hallsworth 2015 | UK | Controlled | NAFLD | 29 | NR | 12 | 15:14 | HIIT | Standard care |
| Haufe 2021 | Germany | Controlled | MetS | 314 | 14 | 26 | 160:154 | MICT | Wait-list control |
| Houghton 2017 | UK | Controlled | NASH | 24 | NR | 12 | 12::12 | MICT + RT | Standard care |
| Kelardeh 2020 | Iran | Placebo | NAFLD | 45 | 100 | 12 | 23:22 | a) RT<br>b) RT + curcumin | a) Curcumin placebo<br>b) Curcumin |
| Pugh 2013 | UK | Controlled | NAFLD | 13 | 54 | 16 | 6:5 | MICT | Standard care |
| Pugh 2014 | UK |  | NAFLD | 21 | 48 | 16 | 18:13 | MICT | Diet and PA advice |
| Rezende 2016 | Brazil | Controlled | NAFLD | 44 | 100 | 24 | 21:23 | MICT | Control |
| Sirisunhirun 2022 | Thailand |  | Liver Cirrhosis | 40 | 35 | 12 | 20:20 | MICT + RT | PA advice & self-monitoring |
| St George 2009 | Australia | Controlled | NAFLD | 141 | 38 | 13 | 107:34 | a) MICT + 3 CS<br>b) MICT + 6 CS<br>c) MICT + 6 CS + TS | Control |
| Sullivan 2012 | USA | Controlled | NAFLD | 18 | 72 | 16 | 12:6 | MICT | Control |
| Takahashi 2015 | Japan |  | NAFLD | 53 | 64 | 12 | 31:22 | RT | Diet and PA advice |
| Tutino 2018 | Italy | Comparative | NAFLD | 142 | 40 | 13 | 101:41 | a) MICT<br>b) MICT + RT c) MICT + LGIMD<br>d) MICT + RT +<br>LGIMD | a) Control diet<br>b) LGIMD |
| Zelber-Sagi<br>2014 | Israel | Placebo | NAFLD | 82 | 47 | 12 | 44:38 | RT | Stretching |
| Zhang 2016 | China | Controlled | NAFLD | 220 | 68 | 48 | 146:74 | VICT + MICT | Control |

The type of liver disease studied included NAFLD (animal = 17, human = 14), NASH (animal = 5, human = 1), hepatic steatosis (animal = 4), liver cirrhosis (human = 1), liver fibrosis (animal = 1), polycystic liver disease (animal = 1), metabolic syndrome (human = 1), type II diabetes (human = 1), and extrahepatic obstruction (animal = 1).

All but one (24) of the human studies provided details on how NAFLD was diagnosed. NAFLD diagnosis methods in human studies included: proton magnetic resonance spectroscopy (MRS) alone (25–27), ultrasonography alone (22), MRS and ultrasonography (28), ultrasonography and elevated alanine aminotransferase (ALT) (29), Asia Pacific Working Party guidelines (30,31), liver biopsy (23), and controlled attenuation parameter (CAP) measurements performed via vibration-controlled elastography (VCTE) (32). Liver biopsies were used to diagnose NASH in the only study that consisted of patients diagnosed with this condition (33). Whereas, in one study (34) a variety of measures were used to diagnose liver cirrhosis (computed tomography, MRI, transient elastography, or liver biopsy).

Human studies had one (N = 13), two (N = 4), or three (N = 1) intervention arms that were relevant to this review. The interventions included only moderate-intensity continuous training (N = 8), resistance training (N = 4), high-intensity interval training (N = 2), moderate-intensity continuous training combined with resistance training (N = 4), vigorous-intensity training (N = 1), or physical activity guidelines and counselling support (N = 1). The comparison conditions included a no intervention control (N = 7), standard care (N = 3), general diet and/or physical activity advice (N = 3), a wait-list control (N = 1), placebo supplement (N = 1), control diet (N = 1), or stretching (N = 1).

Study characteristics of animal studies are presented in Table 2. Animal studies included an approximate total of 992 animals and had a median [inter-quartile range (IQR)] of 32 (24–39), although it was not possible to discern the exact number due to study reporting quality (e.g., a study may have reported ‘7–8’ mice per group). There were 17 studies with rats and 12 studies with mice.

In animal studies, these diseases were induced via diet (N = 22), genetic phenotype (N = 6), or drug induced (N = 1). Animal studies had either one (N = 17) or two (N = 12) intervention arms relevant to this review. Interventions in animal studies included continuous exercise (e.g., treadmill running, N = 24), voluntary exercise (e.g., voluntary wheel running, N = 8), or swimming (N = 5).

Only one study (human = 1) directly reported the number of NAFLD participants who experienced disease regression, reduction, or progression. In most studies, liver function and/or damage were used as surrogate markers for remission and progression, and included tests for liver enzymes: alanine transaminase (ALT) (animal = 18, human = 16), aspartate transaminase (AST) (animal = 11, human = 15), alkaline phosphatase (ALP) (animal = 2, human = 4), and gamma-glutamyl transferase (GGT) (animal = 1, human = 12).

The assessment of liver triglyceride content included hepatic triglyceride content (HTGC) (animal = 9), intrahepatic triglyceride content (IHTG) (human = 1), HTGC/IHTG (human = 5), liver fat percentage (human = 3), intra-hepatocellular lipid (IHCL) (human = 1), liver lipid percentage (animal = 1), intrahepatic lipid content (IHL) (human = 4), liver triacylglycerol (TAG) (animal = 5), and hepatic fat content (HFC) (human = 1).

For the evaluation of liver steatosis and related measures, the fatty liver index (human = 1), ferritin (human = 4), fetuin A (human = 1), hepatic steatosis (human = 2), NAFLD (non-alcoholic fatty liver disease) (human = 1), NAFLD activity score (NAS) (animal = 5), fibrosis score (animal = 1, human = 2), and steatosis score (animal = 3) were included.

Additionally, other liver outcome measures were taken into account, such as liver droplet size (animal = 4), liver stiffness (human = 2), spleen stiffness (human = 1), liver diacylglycerol (DAG) (animal = 3), hepatorenal index (HRI) (human = 1), liver platelets (PLT) (human = 1), hepatic tuberculosis (TB) (human = 1), liver weight (animal = 6), and vascular endothelial growth factor (VEGF) (human = 1).

### Liver disease regression and/or progression

In our review, only one study (25) directly investigated the impact of exercise on the remission or progression of NAFLD. The findings revealed that 14% (n = 3/22) of participants assigned to the exercise group achieved a decrease in hepatic fat content (HFC) to less than 5.6% (normal levels), while this was observed in only 11% (n = 2/18) of those who received no intervention. Moreover, 54% (n = 12/22) of the exercise group experienced a reduction in HFC, while only 17% (n = 3/18) in the no intervention group showed a similar improvement. Furthermore, elevated HFC was observed in only 32% (n = 7/22) of participants in the exercise group, compared to 72% (n = 13/18) in the no intervention arm. These findings suggest a positive association between exercise intervention and the regression of early fibrosis, as well as a delay in the progression to cirrhosis. We found no studies that directly evaluated the progression to HCC.

### Markers of liver function and damage

Meta-analysis of human studies was possible for outcomes ALT, AST, ALP, GGT, and hepatic triglyceride content only (Figure 2). Meta-analysis of animal studies was possible for outcomes ALT, AST, NAS, and liver triglycerides only (Figure 3). We conducted separate meta-regression analyses for human (Supplementary Table 1) and animal studies (Supplementary Table 2). In human studies, we accounted for year, region, disease, and study type. Meanwhile, in animal studies, we controlled for year, region, animal type, disease, and inducement. Results of meta-regression and funnel plots are presented in supplementary material. Results of individual studies not included in the meta-analyses are presented in supplementary table 3 and 4.

### Liver enzymes

Meta-analysis of studies that assessed the effects of exercise on ALT are presented in Figure 2A (human studies) and Figure 3A (animal studies). In the human studies, participants who were assigned to the exercise group exhibited a modest reduction in ALT levels compared to those in the control group (Studies = 12, N = 1,209, SMD = -0.28, 95%CI = -0.53, -0.03). Moreover, in animal studies, a greater difference in ALT levels was observed between the exercise and control groups (Studies = 15, N = 274, SMD = -2.61, 95%CI = -3.91, -1.30). Heterogeneity was considerable in animal studies (I^2^ = 94%) and substantial in human studies (I^2^ = 65%). Some of the heterogeneity in the animal studies could be explained by animal type, with higher effect estimates observed for studies with rats compared to mice (Supplementary Table 2). Funnel plots suggested likely publication bias in animal studies but not for human studies (Supplementary Figure 1). Additional individual studies not included in the meta-analysis reported either a statistically significant reduction in ALT following intervention compared to the control group (35) or no statistically significant difference (28,36) (Supplementary Table 3).

Meta-analysis of studies that assessed the effects of exercise on AST are presented in Figure 2B (human studies) and Figure 3B (animal studies). Participants assigned to the intervention showed a slight difference in AST compared to the control group, favouring the intervention (Studies = 10, N = 1,148, SMD = -0.12, 95%CI = -0.32 to 0.07). However, the certainty of this finding was limited by confidence intervals that crossed the null. Additionally, animals assigned to the exercise intervention demonstrated a significant difference in AST compared to the control group (Studies = 8, N = 146, SMD = -2.88, 95%CI = -4.78 to -0.98). Heterogeneity was considerable in animal studies (I^2^ = 95%) and moderate in human studies (I^2^ = 39%). Funnel plots suggested likely publication bias in animal studies but not in human studies. Meta-regression results are presented for human (Supplementary Table 1) and animal studies (Supplementary Table 2) in the supplementary materials. Some of the heterogeneity in the human and animal studies meta-analysis was explained by study region. Furthermore, in the animal studies, study year explained some of the heterogeneity in the meta-analysis. Individual studies that were not included in the meta-analysis identified a statistically significant reduction in AST in the intervention compared to control arm following intervention (35) or no statistically significant difference (28,36) (Supplementary Table 3).

Meta-analysis of human studies that assessed the effects of exercise on ALP are presented in Figure 2C. Despite the confidence intervals crossing the null, our analysis revealed a small increase in ALP among those assigned to exercise when compared with the control group (Studies = 3, N = 130, SMD = 0.23, 95% CI = -0.13, 0.59). There was no evidence of heterogeneity (I^2^ = 0%), nor was there evidence of publication bias. Meta-regression was not possible for ALP in either human or animal studies. Two individual animal studies examined the effects of exercise on ALP, both found a decreases in ALP for exercise compared to control (37,38) (Supplementary Table 4).

Meta-analysis of human studies that assessed the effects of exercise on GGT are presented in Figure 2D. There was a small decrease in GGT in participants receiving exercise intervention compared to controls (Studies = 10, N = 1,142, SMD = -0.23, 95%CI = -0.36, -0.10). Heterogeneity was low (I^2^ = 6%) and there was no clear evidence of publication bias (Supplementary Figure 1). There were no statistically significant findings from the meta-regression (Supplementary Table 1). In three individual studies not included in the meta-analysis, two (35,36) identified decreases in GGT following exercise relative to control (Supplementary table 3). One study (28) did not identify definitive differences between exercise and control groups. There was no difference in GGT between groups in one animal study (39) (Supplementary Table 4).

### Liver triglyceride content

Meta-analysis of human studies that assessed the effects of exercise on HTGC are presented in Figure 2E. Participants assigned to exercise showed, on average, HTGC compared to the control group (Studies = 3, N = 101, SMD = -0.24, 95%CI = -0.66, 0.18). However, it’s worth noting that the confidence intervals overlapped the null value. Heterogeneity was low (I^2^ = 0%) and there was no clear evidence of publication bias (Supplementary Figure 1). Meta-regression was not possible for HTGC in human studies. The percentage decrease in HTGC was greater following moderate and high intensity exercise compared to control in one individual human study not included in the meta-analysis (28). There was no difference between moderate and high intensity exercise.

Meta-analysis of animal studies that assessed the effects of exercise on liver triglycerides are presented in Figure 3D. There was a large difference in liver triglycerides between exercise and control groups (Studies = 10, N = 172, SMD = -1.94, 95%CI = -3.87, 0.00). Heterogeneity was, however, considerable (I^2^ = 96%) and there was evidence of publication bias (Supplementary Figure 2). Meta-regression indicated that some of the heterogeneity could be explained by animal type—with larger effect estimates observed for studies that used rats—but not by year, region, disease, or inducement (Supplementary Table 2). Individual animal studies not included in the meta-analysis suggested lower liver triglycerides for exercise compared to control (40).

For outcomes not included in the meta-analysis (Supplementary Table 3 and Table 4), individual studies suggested that exercise can decrease liver TAG (41–43), HFC (25), the fatty liver index (44), IHCL (45), and IHL (24,46).

### Liver steatosis and related measures

Meta-analysis of all four animal studies that assessed the effects of exercise on NAS are presented in Figure 3C. There was a large decrease in NAS with exercise versus control (Studies = 4, N = 98, SMD = -1.36, 95%CI = -2.08, -0.65). Heterogeneity was substantial (I^2^ = 59%) and there was some evidence of publication bias (Supplementary Figure 2). Meta-regression of NAS was not possible and therefore, the heterogeneity observed remained unexplained.

For outcomes not included in the meta-analysis (Supplementary Table 3 and Table 4), the findings on Ferritin were mixed in the literature; two studies (35,36) reported a decrease with exercise compared to controls, while two other studies (23,31) found no statistically significant difference. Similarly, one study (47) observed differences in fibrosis score between exercise and control groups, while another (33) did not find any distinction.

### Other relevant liver outcome measures

Meta-analysis of all six animal studies that assessed the effects of exercise on liver weight are presented in Figure 3E. There was a large difference in liver weight between exercise and control groups (Studies = 6, N = 144, SMD = -1.94, 95%CI = -2.78, -1.10). Heterogeneity was considerable (I^2^ = 77%), although could not clearly be explained by factors included in the meta-regression analysis (Supplementary Table 2). There was possible publication bias evident in the funnel plots (Supplementary Figure 2).

For outcomes not included in the meta-analysis (Supplementary Table 3 and Table 4), individual studies suggested that exercise can decrease liver DAG (42,48), hepatic stiffness (31) and HRI (36). There were null findings for liver stiffness (34), spleen stiffness (34), and VEGF (23).

## Discussion

This current review synthesized the evidence for the effect of exercise interventions versus control/standard care on liver function and disease in people and animals with established liver disease. Only one study directly reported whether liver disease (NAFLD) regressed or progressed in response to exercise intervention. Most studies included in this review compared measures of liver function and/or damage between exercise and non-exercise interventions. The findings of these studies support the biological plausibility for a protective effect of exercise on liver function and disease.

The primary strength of this review is the synthesis of findings from both human and animal studies. While there is a wealth of data from animal studies relevant to human health, systematic reviews, which are common practice in clinical and public health research, remain less common in animal research. Animal studies offer a unique advantage in exploring liver outcome measures that are not feasible to obtain through biopsy in humans. However, it is essential to recognize that the responses observed in animals may not always directly translate to human-specific outcomes. On the other hand, human studies heavily rely on surrogate measures of liver function and damage. By incorporating results from both human and animal studies, this review greatly enhances our understanding of the effects of exercise on liver function, as well as the potential translation of animal study findings to human contexts.

This current review also has limitations that should be considered when interpreting the findings. First, we did not perform a risk of bias assessment using a pre-existing risk of bias tool. While tools for assessing study quality in human clinical trials have been modified and applied to animal studies (e.g., SYRCLE), when piloting this tool most animal studies scored poorly simply because the information was not reported.

There was substantial to considerable heterogeneity in the meta-analyses, particularly the meta-analyses of the animal studies, which may limit confidence in the final effect estimates. Sources of heterogeneity present in animal studies that were not present in the human studies included species (e.g., mouse type, rat type) as well as methods used to induce liver disease (e.g., forced feeding, genetic selection). These are in addition to sources of variation present in human studies, such as variations in study design and methods, participant age and sex, exercise type, dose, or duration, and outcome assessment methods. Owing to the studies identified, this review is limited in its ability to examine differing effects for the type and dose of exercise on liver function and/or damage.

Our study’s findings indicate that exercise interventions have a positive impact on liver health and function when compared to non-exercise comparison groups. These results align with evidence from epidemiological studies, which have shown that higher levels of physical activity are associated with reduced instances of fatty liver and a decreased risk of liver cancer. For example, in a meta-analysis of observational studies (49), higher (90th percentile) compared to lower (10th percentile) duration of leisure-time physical activity was associated with decreased liver cancer risk (HR = 0.73, 95%CI: 0.55, 0.98). While this epidemiological evidence suggests an association between exercise and outcomes like liver cancer, results from this current review support the biological plausibility of the findings. That is, exercise may reduce the risk of liver cirrhosis and/or cancer by reducing measures of liver function and/or damage.

Improvement in liver enzymes, liver triglyceride content, and steatosis was evident in both human and animal studies, but the effect estimates were generally larger in the meta-analysis of animal studies compared with the human studies. This may, in part, reflect inter-species differences, with greater potential for liver regeneration in animals. Alternatively, these larger effect estimates may be due to a greater ability to control for sources of variation in animal compared to human studies, who are likely to vary in factors such as intervention adherence, diet, alcohol consumption, body fatness, as well as exercise history and tolerance. Nonetheless, that improvements were present in both humans and animals supports the translation of findings and relevance to people with liver disease.

This current review found an effect of exercise on liver outcomes, but it did not attempt to determine whether these are direct or indirect effects. That is, is the benefit of exercise due to the exercise itself or other factors such as fat loss that may follow exercise participation. Findings from individual studies included within the review suggest at least some of the effects of exercise may be via fat loss (28). However, other studies have proposed mechanisms independent of fat loss, such as facilitating change via the effect of exercise on insulin signalling and serum lipid concentrations, which may lead to an increased ability to suppress lipolysis in adipose tissue, resulting in decreased free fatty acid delivery to the liver (47).

The identification of exercise’s impact on liver function and damage provides essential support for the idea that exercise might be linked to liver cancer. As liver cancer is a rare form of cancer, directly examining this relationship presents challenges. Nevertheless, it is important to emphasize that this discovery is just one step in the broader research pathway. Conducting further research is necessary to delve into how these measures specifically influence the incidence of liver cirrhosis and/or cancer (HCC).

## Conclusion

In conclusion, a single investigation focusing on the effect of exercise on the progression and remission of NAFLD revealed a significant association between exercise intervention and the regression of initial fibrosis, as well as a delay in the progression of cirrhosis. Moreover, a comprehensive review of existing literature reveals consistent indications of exercise’s positive impact on liver function and reduction of liver damage in both human subjects and animal models with established liver disease. These collective results substantiate the biological plausibility of exercise as a beneficial therapeutic strategy for enhancing liver function and combatting liver disease. Nonetheless, further research and clinical trials are warranted to fully elucidate the mechanisms underlying these observed effects and to establish more robust guidelines for exercise-based interventions in liver disease management.

## Data Availability

All data produced in the present study are available upon reasonable request to the authors

## Acknowledgments

All authors were involved in conception and design. A. E. D. performed the literature searches, with A. E. D. and J. K. V. selecting articles for exclusion and inclusion, with any disagreement resolved by discussion with B. M. L. Full texts were assessed by E. N. S and I. M. L, with any disagreement resolved by discussion with B. M. L. Data was extracted from human studies by E. N. S. and I. M. L, and from animal studies by A. E. D. and B. M. L. Meta-analysis was conducted by C. T. V. S. All authors contributed to the writing and revision of the manuscript.

---

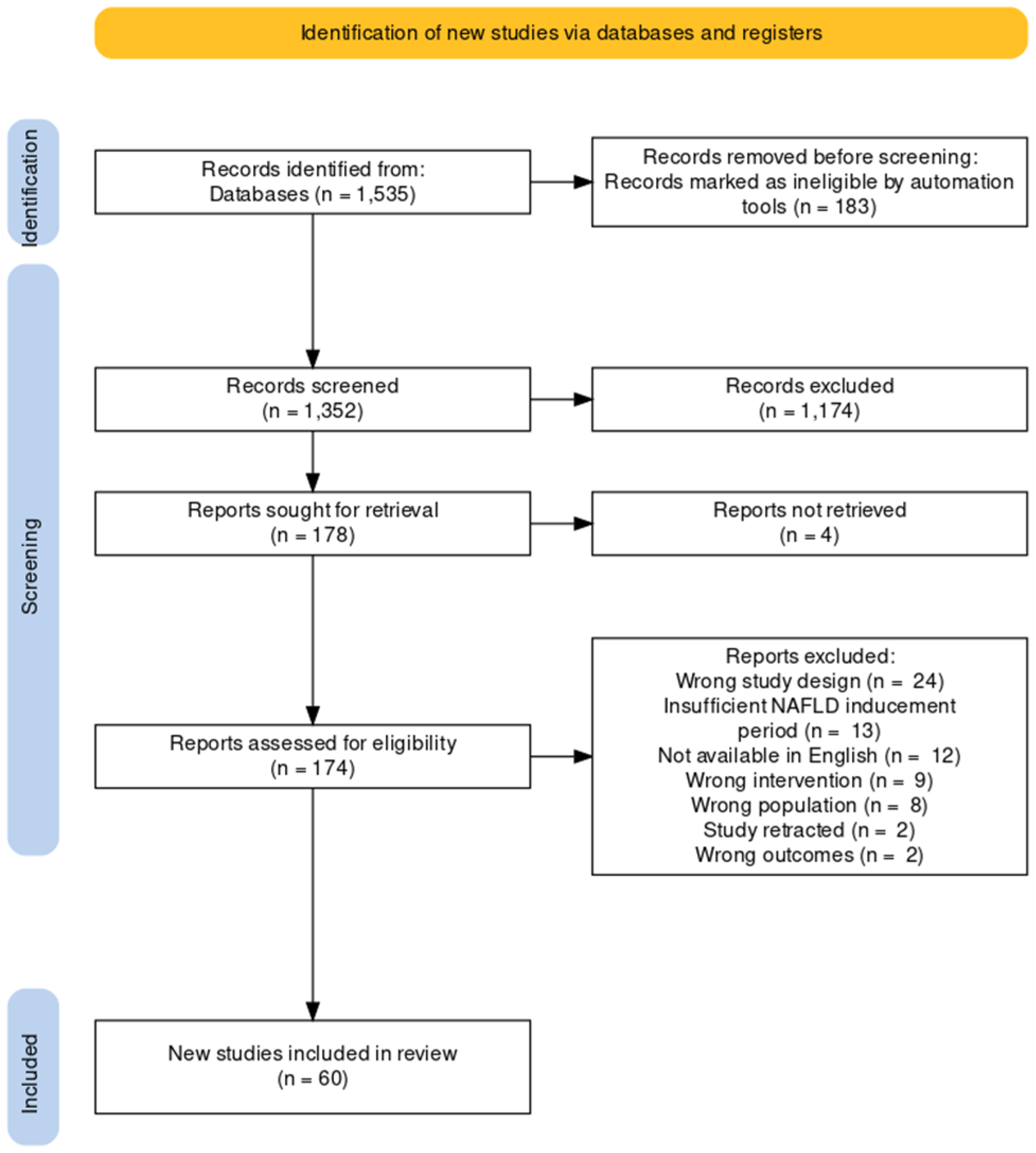

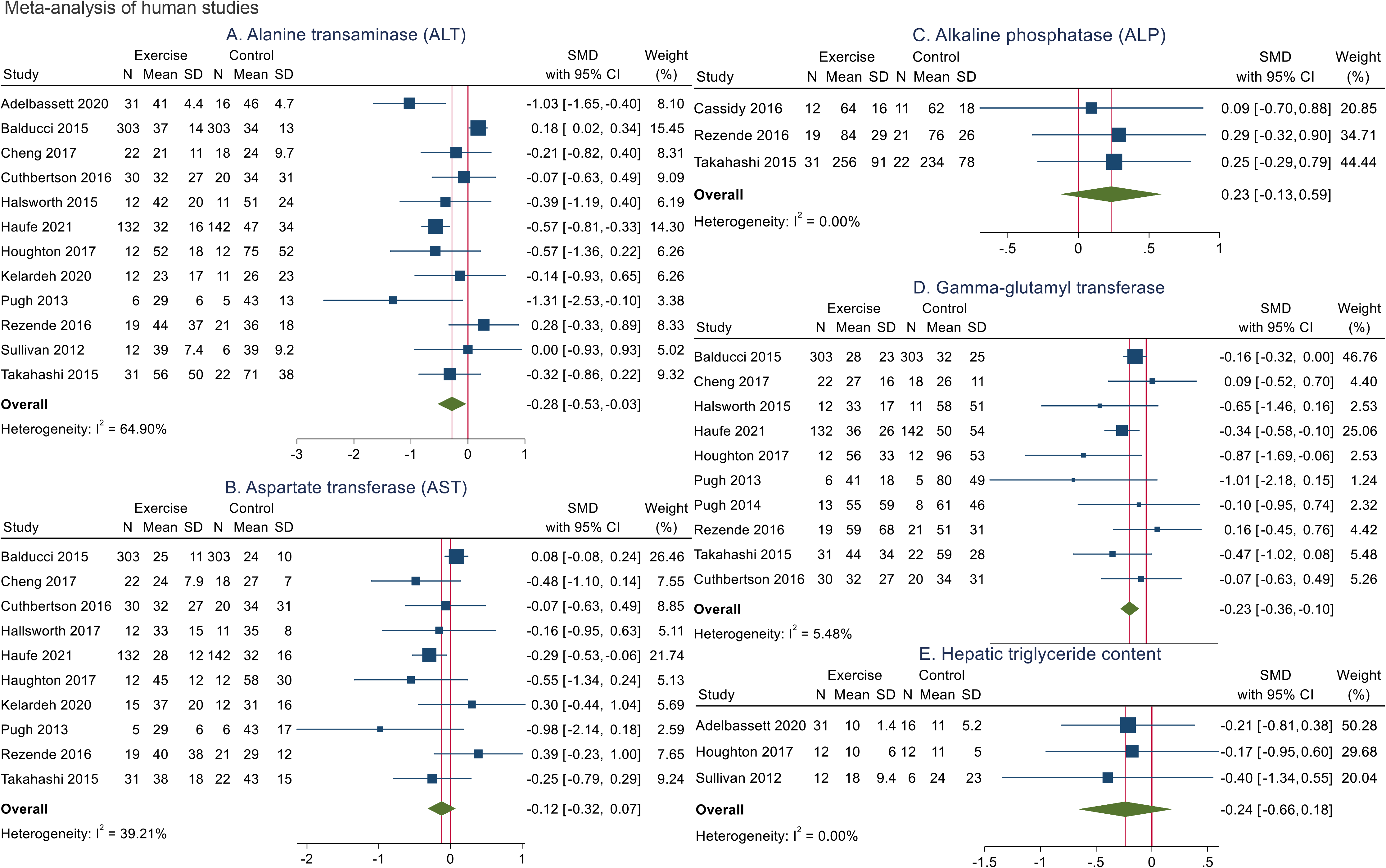

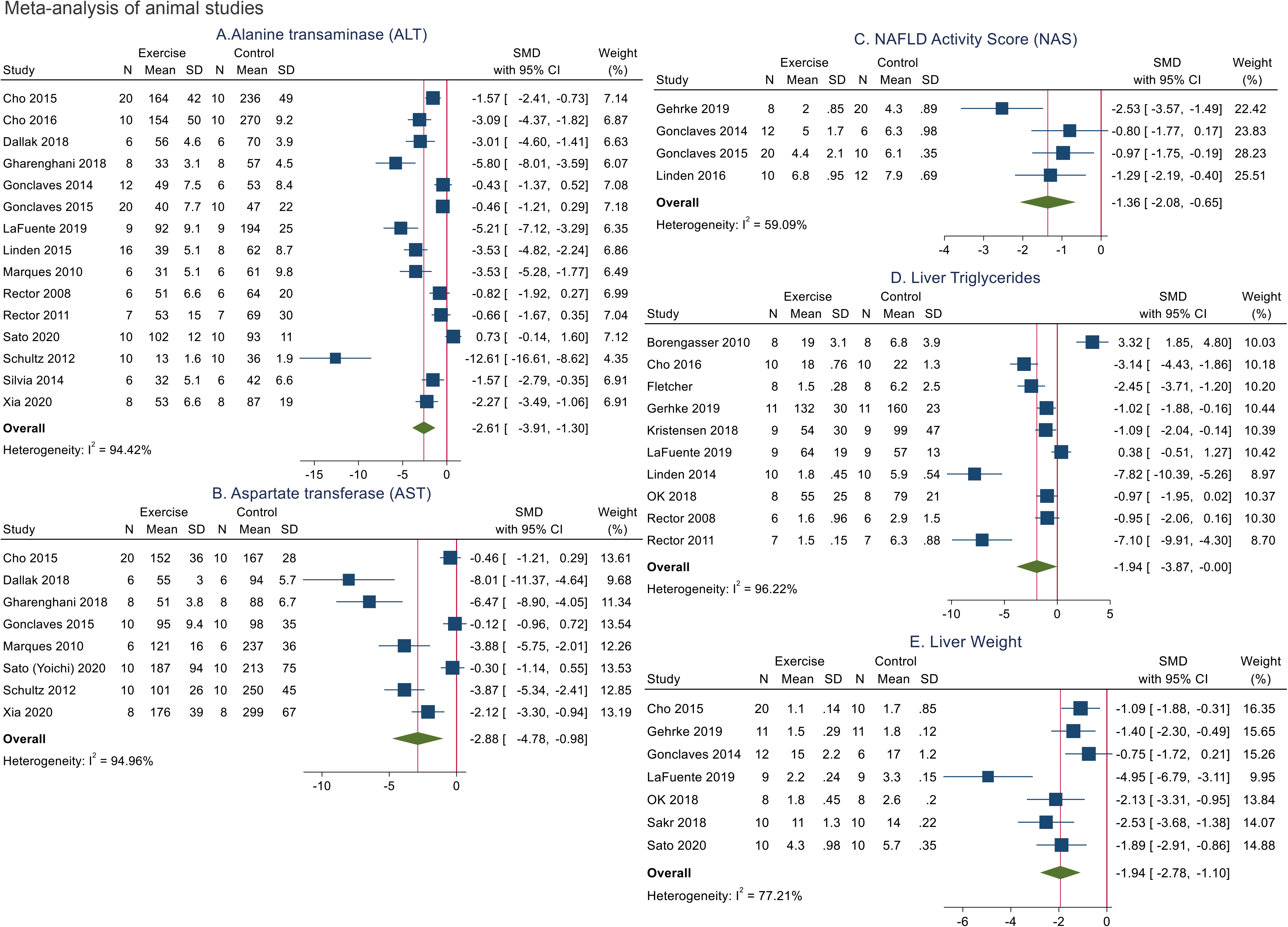

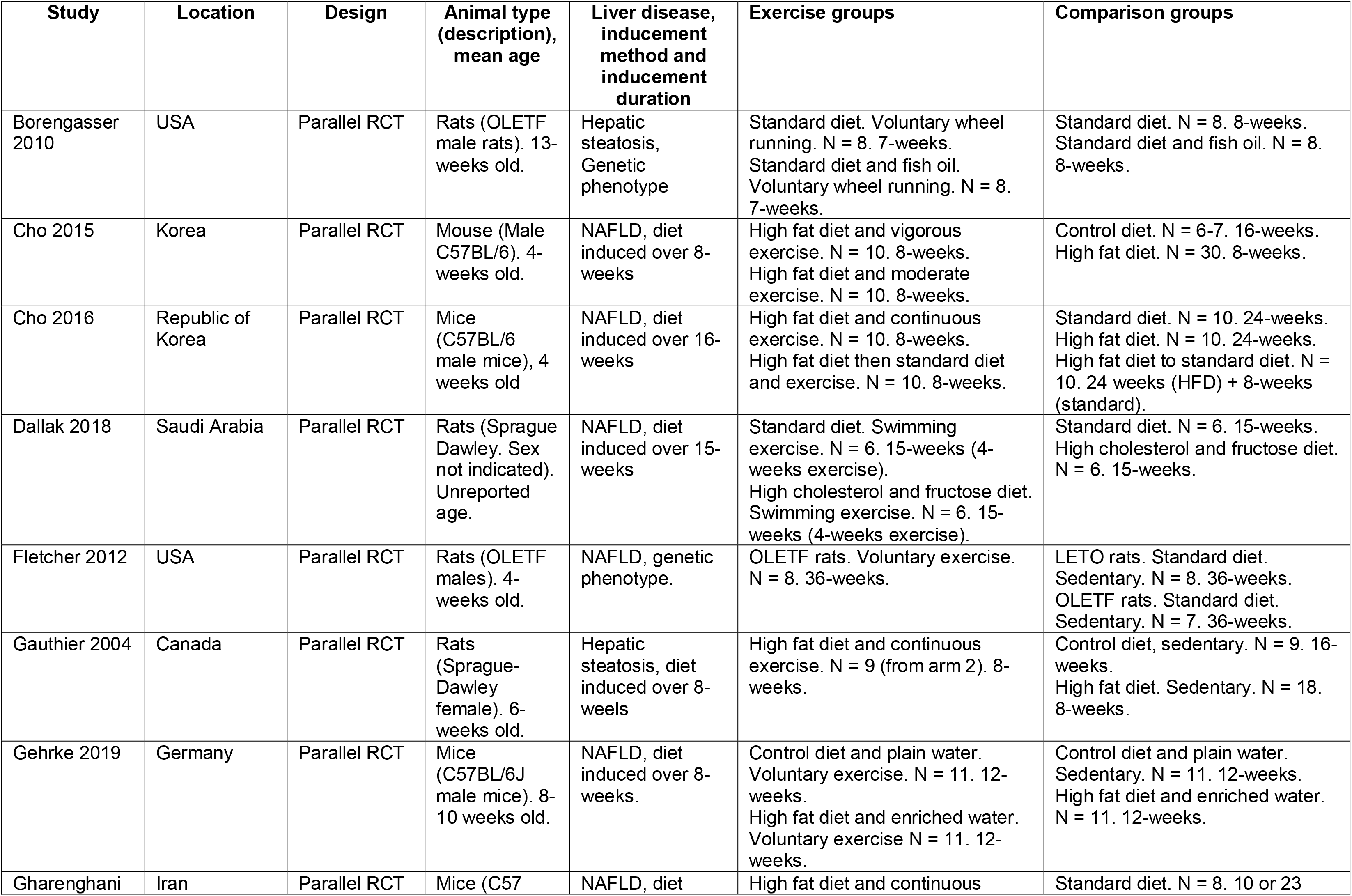
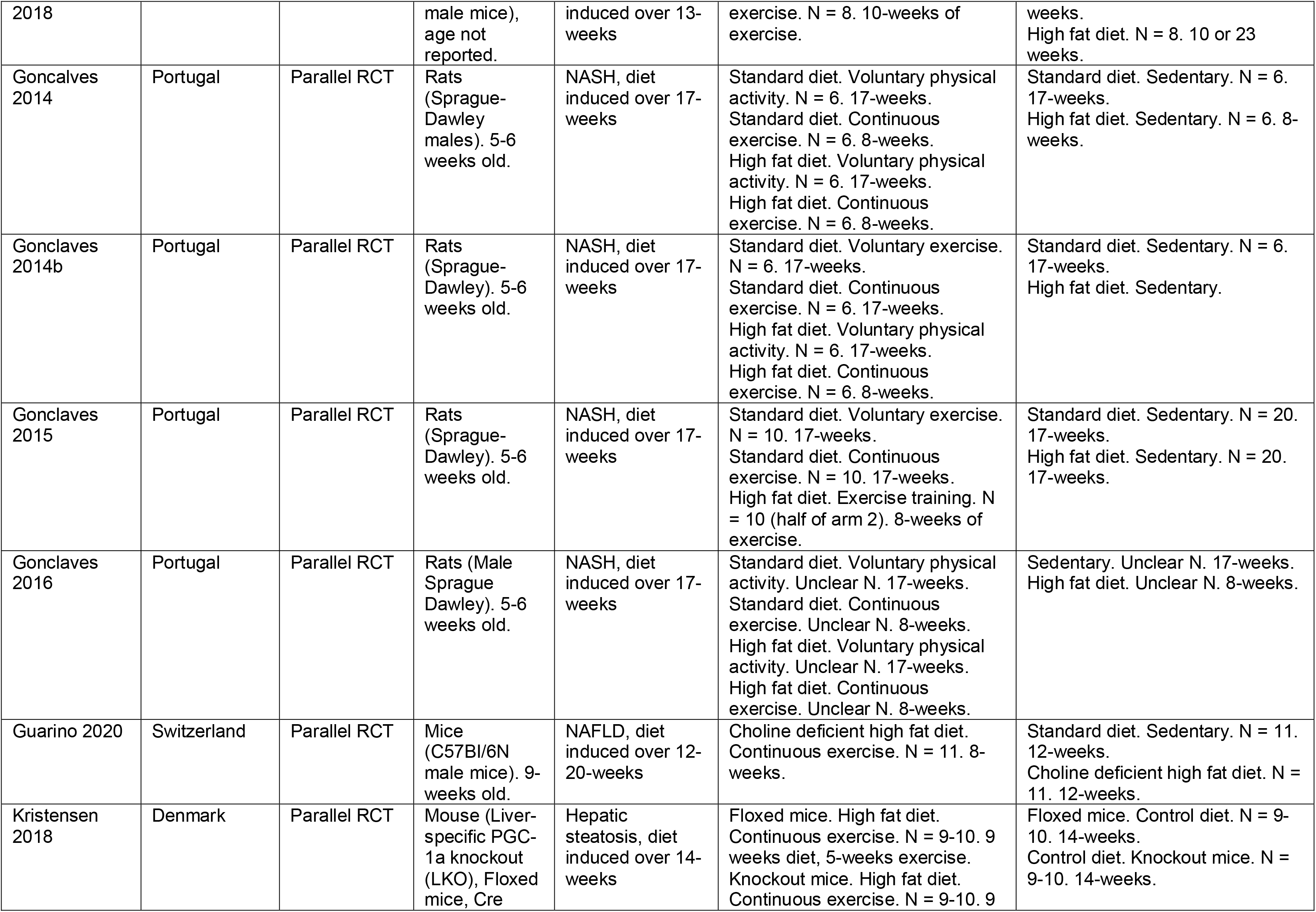
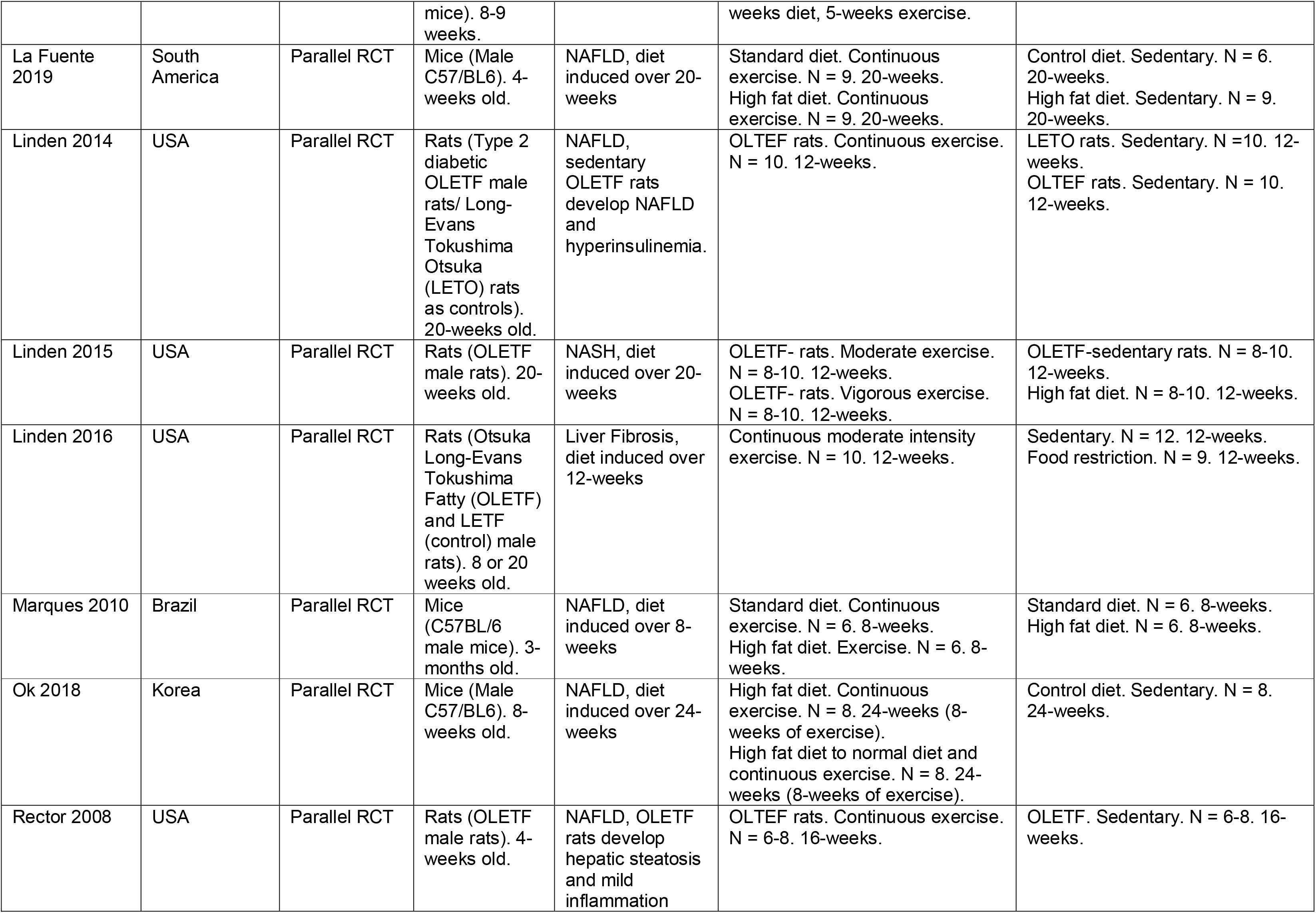
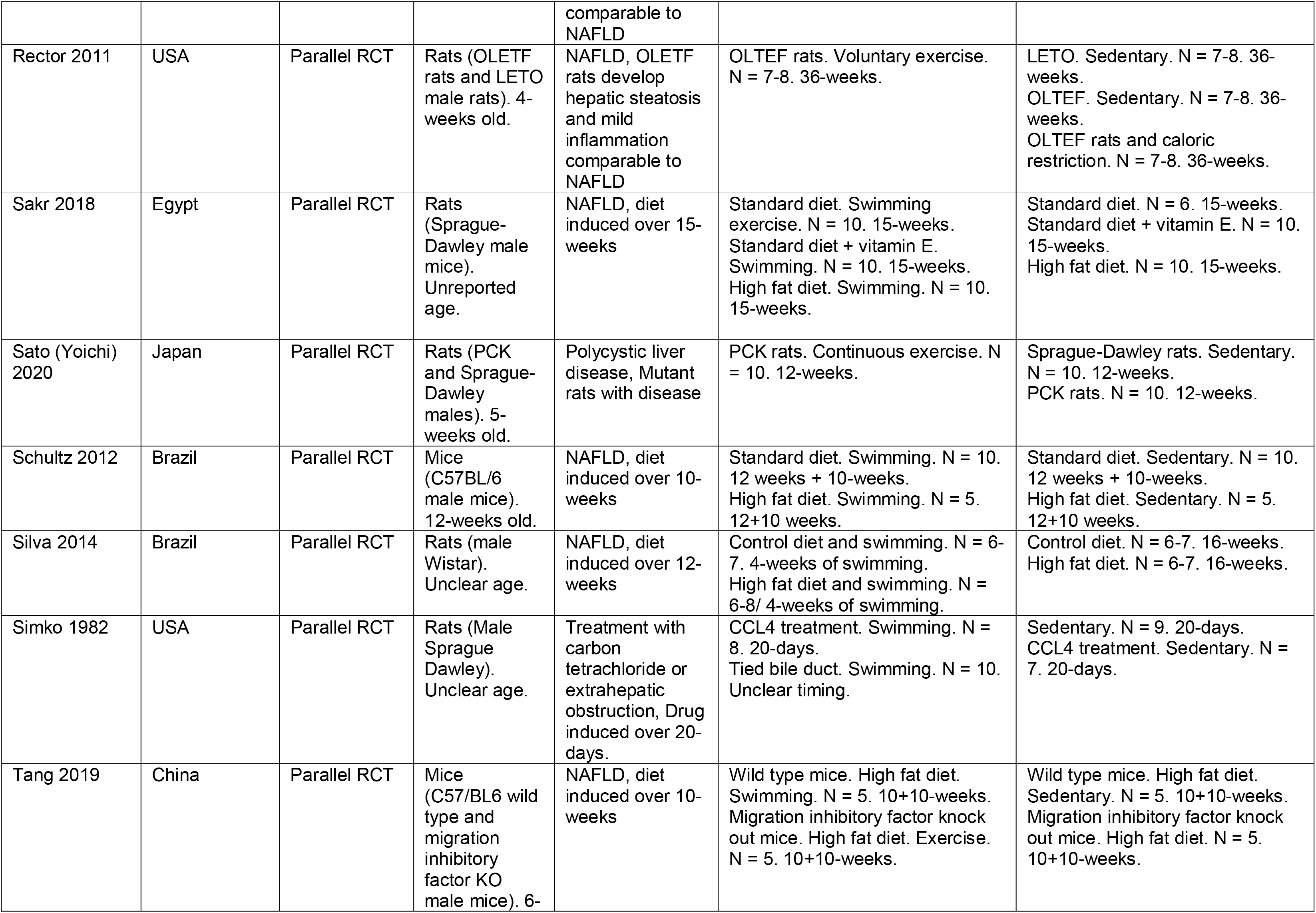
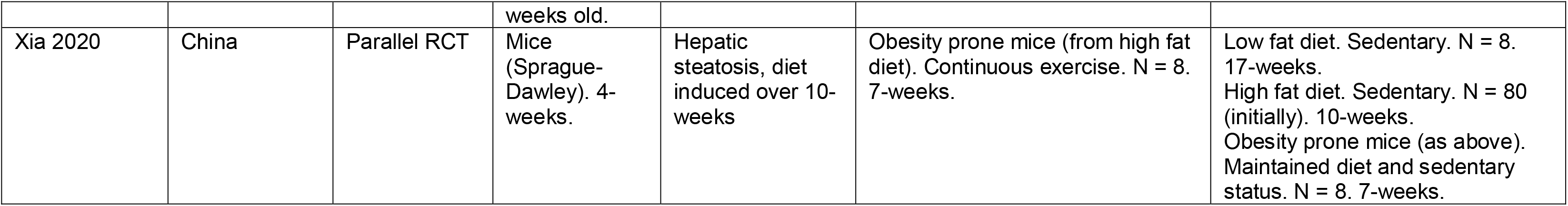

