## Supplementary Materials for "The role of exercise in limiting progression from liver inflammation and fibrosis to cirrhosis and carcinoma: a systematic review with meta-analysis of human and animal studies"

**Supplements**

**Search strategy**

Medline Search June 2021

1. exp Water Sports/ or exp Racquet Sports/ or exp Sports for Persons with Disabilities/ or exp Youth Sports/ or exp Snow Sports/ or Sports/ or exp Exercise Movement Techniques/ or exp Warm-Up Exercise/ or exp Exercise/ or exp Exercise Therapy/ or exp Circuit-Based Exercise/ or exp Cool-Down Exercise/ or exp Plyometric Exercise/

2. exp High-Intensity Interval Training/ or exp Bicycling/ or exp dancing/ or exp resistance training/ or exp football/ or exp soccer/ or exp jogging/ or exp tennis/

3. exp Gymnastics/ or exp Athletic Performance/ or exp Swimming/ or exp Running/ or exp Boxing/ or exp Weight Lifting/ or exp physical fitness/ or exp Walking/ or exp Stair Climbing/

4. (exercise or dancing or circuit or running or cycling or plyometric or golf or gymnastics or resistance or jogging or boxing or football or volleyball or wrestling or skiing or water sports or bicycling or snow sports or skating or martial arts or hockey or baseball or mountaineering).ti,ab,kf.

5. (sport* or physical fitness or strengthening or basketball or weight lifting or soccer or skating or walking or bowling or athletic or diving or stair climbing or yoga or pilates or racquet or muscle stretching exercise* or physical conditioning or high intensity interval training or endurance training).ti,ab,kf.

6. (physical activit* or physical inactivit* or interval training or exertion or workout).ti,ab,kf.

7. 1 or 2 or 3 or 4 or 5 or 6

8. Fatty Liver/

9. Liver Diseases/

10. Liver Cirrhosis/

11. (fatty liver or liver fibrosis or liver cirrhosis or liver disease*).ti,ab,kf.

12. (Non-alcoholic steatohepatitis or steatohepatitis or alcoholic fatty liver disease or non-alcoholic fatty liver disease or steatosis or intrahepatic lipid*).ti,ab,kf.

13. Hepatitis, Alcoholic/ or Hepatitis, Chronic/ or Hepatitis B, Chronic/ or Hepatitis D, Chronic/ or Hepatitis, Autoimmune/ or Hepatitis, Viral, Human/ or Hepatitis C, Chronic/ or Hepatitis/

14. hepatitis.ti,ab,kf.

15. 8 or 9 or 10 or 11 or 12 or 13 or 14

16. Physical Conditioning, Human/

17. physical conditioning.ti,kf,ab.

18. 16 or 17

19. Physical Conditioning, Animal/

20. wheel running.ti,kf,ab.

21. 19 or 20

22. 18 or 21

23. 7 or 22

24. 15 and 23

25. 24 and "Animals".sa_suba.

26. 24 and "Humans".sa_suba.

27. 25 or 26

28. 27 and "Fatty Liver".sa_suba.

29. limit 28 to english language

30. limit 29 to "review articles"

31. 29 not 30

32. (exercise or physical activit*).ti,ab,kf.

33. 31 and 32

34. limit 24 to english language

35. limit 34 to "review articles"

36. 34 not 35

37. 32 and 36

38. exp Fatty Liver/

39. 37 and 38

**Table 1: Meta-regression of human studies**

| **Factor** | **B (95% CI)** |
| --- | --- |
| **ALT** | |
| Year | -0.06 (-0.17, 0.5) |
| Region | 0.08 (-0.19, 0.36) |
| Disease | 0.01 (-0.29, 0.32) |
| Type | 0.05 (-0.20, 0.30) |
| **AST** | |
| Year | -0.02 (-0.10, 0.07) |
| Region | 0.28 (0.02, 0.56) |
| Disease | -0.04 (-0.29, 0.21) |
| Type | 0.08 (-0.20, 0.30) |
| **GGT** | |
| Year | 0.02 (-0.10, 0.14) |
| Region | 0.06 (-0.26, 0.37) |
| Disease | -0.18 (-0.46, 0.11) |
| Type | -0.11 (-0.39, 0.15) |

**Table 2: Meta-regression of animal studies**

Meta-regression of animal studies

| **Factor** | **B (95% CI)** |
| --- | --- |
| **ALT** | |
| Year | -0.15 (-0.43, 0.14) |
| Region | -0.85 (-1.4, -0.26) |
| Animal | 2.49 (0.79, 4.20) |
| Disease | 0.21 (-0.85, 1.28) |
| Inducement | 1.01 (-1.00, 3.03) |
| **AST** | |
| Year | -0.82 (-1.32, -0.32) |
| Region | -2.25 (-3.35, -1.17) |
| Animal | 1.24 (-0.74, 3.22) |
| Disease | 0.09 (-2.03, 2.21) |
| Inducement | 2.39 (-3.30, 8.08) |
| **Liver Triglycerides** | |
| Year | -0.38 (-2.02, 1.26) |
| Region | 1.06 (-1.48, 3.60) |
| Animal | -14.46 (-26.62, -2.30) |
| Disease | 3.11 (-1.46, 7.69) |
| Inducement | 9.86 (-3.45, 23.16) |
| **Liver Weight** | |
| Year | 0.03 (-1.41, 1.46) |
| Region | -1.48 (-4.47, 1.51) |
| Animal | 3.14 (-4.50, 10.77) |
| Disease | -2.77 (-14.68, 9.14) |
| Inducement | 1.69 (-13.33, 16.72) |

**Figure 1: Funnel plots for the meta-analyses of human studies**

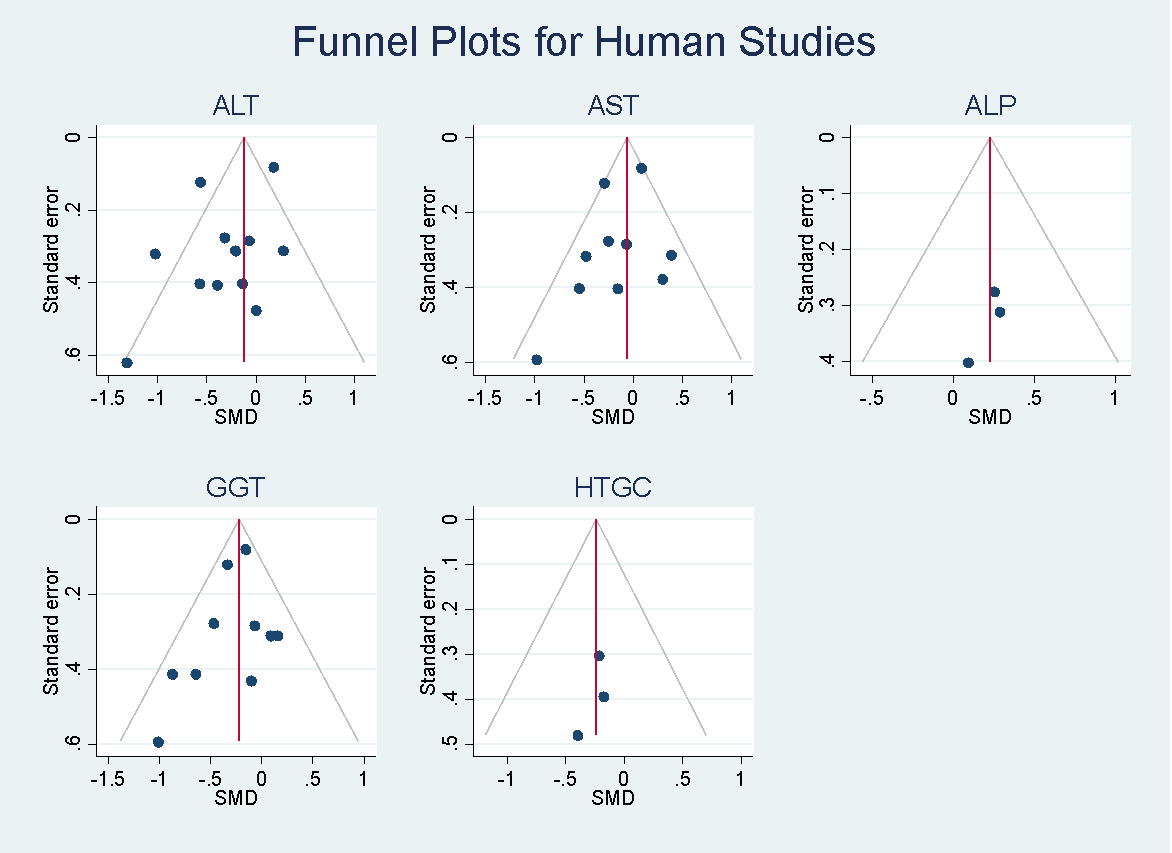

**Figure 2: Funnel plots for the meta-analyses of animal studies**

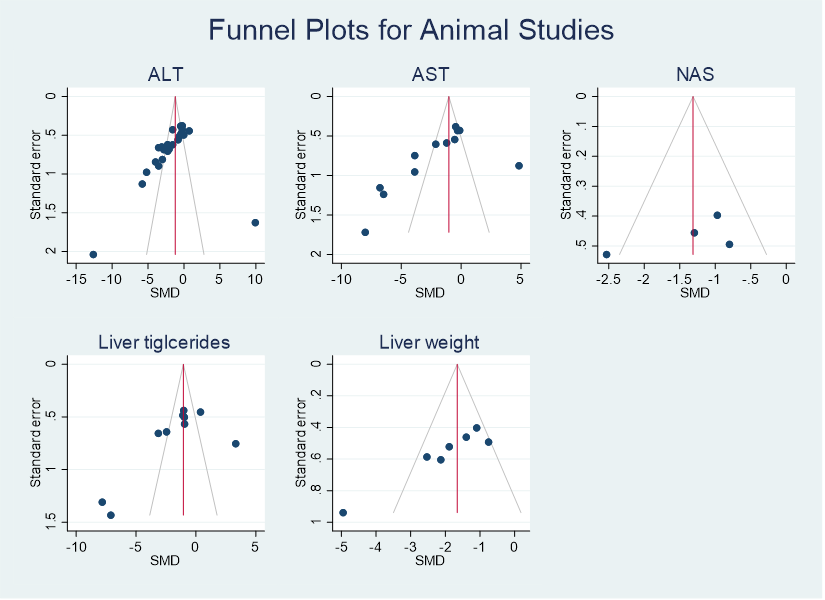

**Table 3: Results from human studies not included in the meta-analysis.**

| **Outcome** | **Study** | **Result** |
| --- | --- | --- |
| **ALT** | St George 2009 | Decreased following increase in physical activity compared to remaining sedentary. Mean (SD) change following increase in physical activity = -14.8 (26.0) compared to -2.4 (29.2) for remaining sedentary. |
|  | Zelber-Sagi 2014 | No difference in responses decrease for the resistance group compared to stretching (P = 0.95). Mean (SD) change was -5.30 (9.65) for the resistance group and -5.10 (14.43) for the stretching group. |
|  | Zhang 2016 | No definitive differences between groups. Percentage change at 12-months was -1.3 (-3.8 to 1.3) for the control, -2.2 (-4.8 to 0.4) for the moderate intensity, and -2.7 (-5.3 to -0.01) for the high intensity exercise group. |
| **AST** | St George 2009 | Decreased following increase in physical activity compared to remaining sedentary. Mean (SD) change following increase in physical activity = -7.6 (17.8) compared to 2.2 (17.4) for remaining sedentary. |
|  | Zelber-Sagi 2014 | No difference in responses decrease for the resistance group compared to stretching (P = 0.97). Mean (SD) change was -2.76 (7.75) for the resistance group and -2.68 (6.95) for the stretching group. |
|  | Zhang 2016 | No definitive differences between groups. Percentage change at 12-months was 0.3 (-1.0 to 1.6) for the control, -0.9 (-2.3 to 0.4) for the moderate intensity, and 0.8 (-0.6 to 2.2) for the high intensity exercise group. |
| **ALP** | Kelardeh 2020 | Unable to discern findings due to formatting error in Table 4. |
| **GGT** | St George 2009 | Decreased following increase in physical activity compared to remaining sedentary. Mean (SD) change following increase in physical activity = -17.1 (36.3) compared to 1.0 (27.5) for remaining sedentary. |
|  | Zelber-Sagi 2014 | Greater but non-significant decrease in the resistance group compared to stretching (P = 0.08). Mean (SD) change was -4.25 (13.03) for the resistance group and 2.35 (16.48) for the stretching group. |
|  | Zhang 2016 | No definitive differences between groups. Percentage change at 12-months was -1.7 (-5.2 to 1.9) for the control, -0.4 (-4.0 to 3.2) for the moderate intensity, and -2.7 (-6.4 to 1.0) for the high intensity exercise group. |
| **HTGC** | Zhang 2016 | Percentage decrease in IHTG following moderate and high intensity exercise compared to control. Effect estimates = -3.5 (-5.6 to -1.3) for moderate exercise vs control and -3.9 (-6.0 to -1.7) for high intensity exercise vs control. No difference between moderate and high intensity exercise, effect = -0.4 (-2.6 to 1.8) |
| **Fatty liver index** | Balducci 2015 | Decrease in exercise compared to control. Mean difference (95%CI) = -6.19 (-8.43 to - 3.96). |
| **Ferritin** | Rezende 2016 | No change vs baseline in control (p = 0.83) but non-significant decrease following exercise (p = 0.15). Final mean (SD) = 233.91 (187) for control and 163.24 (191) for the exercise group. |
|  | St George 2009 | Decreased following increase in physical activity compared to remaining sedentary. Mean (SD) change following increase in physical activity = -25.9 (81.5) compared to -8.1 (63.7) for remaining sedentary. |
|  | Takahashi 2015 | No change in intervention compared to control (p = 0.63). Post intervention mean (SD) = 164.6 (110) for control and 174.8 (202.5) for exercise group. |
|  | Zelber-Sagi 2014 | Greater decrease in the resistance group compared to stretching (P < 0.05). Mean (SD) change was -18.29 (48.63) for the resistance group and 8.25 (51.09) for the stretching group. |
| **Fibrosis score** | Houghton 2017 | No time vs treatment interaction and no clear change from baseline for either control or intervention groups. Final mean (SD) for control = -0.98 (1.53), intervention = -1.50 (1.12). |
|  | Haufe 2021 | Decrease following exercise but not control condition. Final mean (SEM) = 0.40 (0.02) for control and 0.35 (0.01) for exercise groups. |
| **Hepatic stiffness** | Takahashi 2015 | Decreased following intervention compared to control (p < 0.01). Post intervention mean (SD) = 2.05 (0.58) for control and 1.53 (0.64) for intervention groups. |
| **HFC** | Cheng 2017 | Aerobic exercise decreased HFC compared to control (p < 0.01). Mean change was -24.4% (95%CI -41.7 to -1 7.1) in the exercise group compared to 20.9% (95%CI -4.4 to 46.2) in the control group. |
| **HRI** | Zelber-Sagi 2014 | Greater decrease in the resistance group compared to stretching (P < 0.05). Mean (SD) change was -0.25 (0.37) for the resistance group and -0.05 (0.28) for the stretching group. |
| **IHCL** | Shojae-Moradie 2016 | Decrease following exercise but not control (p = 0.02). Final median (IQR) values =12.6 (9.2 to 26.1) for control and 8.9 (5.4 to17.3) for exercise. |
| **IHL** | Hallsworth 2015 | Decrease following HIIT compared to control (P = 0.02). Post intervention mean (SD) = 10.4 (3.9) for control and 7.8 (2.4) for intervention group. |
|  | Hallsworth 2011 | Decrease following exercise compared to control (p = 0.05). Post intervention mean (SD) = 11.5 (7.4) for control and 12.2 (9.0) for exercise group. |
| **Liver fat %** | Pugh 2013 | Decrease following exercise but change was non-significant when compared to control (p = 0.18). Post intervention mean (95%CI) = 18.5 (8.4 to 40.9) for the control and 14.2 (5.4 to 37.0) for the intervention group. |
|  | Cassidy 2016 | Decrease following HIIT compared to control. Post intervention mean (SD) = 7.7 (6.9) for control and 4.2 (3.6) for intervention group. |
| **Liver stiffness** | Sirisunhirun 2022 | No difference between intervention and control groups. Mean change (95%CI) = 0.7 (-1.2 to 2.7) |
| **PLT** | Kelardeh 2020 | Unable to discern findings due to formatting error in Table 4. |
| **Spleen stiffness** | Sirisunhirun 2022 | No difference between intervention and control groups. Mean change (95%CI) = 12.3 (-8.7 to 33.3) |
| **TB** | Kelardeh 2020 | Unable to discern findings due to formatting error in Table 4. |
| **VEGF** | Rezende 2016 | No change compared to baseline for control (p = 0.84) or intervention groups (p = 0.8). Final mean (SD) = 269.3 (239.6) for control and 144.3 (156) for exercise group. |

**Table 4: Results from animal studies not included in the meta-analysis.**

| **Outcome** | **Study** | **Finding** |
| --- | --- | --- |
| **ALP** | Marques 2010 | Lower following high fat and exercise (59.3 ± 2.0) compared to high fat sedentary (75.7 ± 0.7). |
|  | Schultz 2012 | Lower following high fat and exercise (23.3 ± 1.0) compared to high fat sedentary (37.5 ± 3.2). |
| **Fibrosis score** | Linden 2016 | Lower following exercise condition (1.9 ± 0.3) compared to the control (2.7 ± 0.2). |
|  | Sato 2020 | Fibrosis index percentage was lower for the exercise condition compared to the sedentary condition. Results were presented graphically. |
| **Liver Steatosis** | Cho 2016 | High fat diet and exercise has a lower steatosis grade (3) than high fat diet alone (4). |
|  | Fletcher 2012 | OLETF exercise had a lower steatosis score (0.7 ± 0.1) than OLETF sedentary rats (2.9 ± 0.1). |
|  | Gehrke 2019 | High fat diet and voluntary exercise (1.0 ± 0) was lower than high fat diet alone (2.4 ± 0.3). |
|  | Rector 2011 | Steatosis was lower following exercise compared to control conditions. |
|  | Schultz 2012 | Steatosis percentage was lower in the high fat and exercise group compared to the high fat only group. Results presented graphically. |
| **Liver DAG** | Linden 2014 | Liver DAGs lower for exercise group. Results presented graphically. |
|  | Rector 2011 | Liver DAGs lower for exercise compared to control conditions. |
| **Liver TAG** | Guathier 2004 | Lower in high fat and trained than high fat alone but SD/SE were not clear. |
|  | Linden 2015 | Lower following both moderate and vigorous exercise compared to sedentary. Results presented graphically. |
|  | Sheldon 2014 | Lower for the exercise (3.3 ± 0.4) compared to sedentary (8.1 ± 1.5) condition. |
| **Liver droplet size** | la Fuente 2019 | Liver droplet size smaller in high fat diet and exercise compared to high fat diet alone. Results presented graphically |
